# Validation of an EEG-based Delirium Monitor in the ICU

**DOI:** 10.64898/2026.09.23.26361767

**Authors:** Chukwudi A. Onyemekwu, Suganya Karunakaran, Michelle Armenta Salas, Niall T. Prendergast, Kelly M. Toth, Melinda D. Miller, Sangil Lee, Baharan Kamousi, Malissa A. Mulkey, Farid Sadaka, Thomas Striegel, Timothy D. Girard

**Affiliations:** Center for Research, Investigation, and Systems Modeling of Acute illness (CRISMA) in the Department of Critical Care Medicine; University of Pittsburgh School of Medicine; Pittsburgh, PA, USA; Ceribell, Inc., Sunnyvale, CA, USA; Division of Pulmonary, Allergy, Critical Care and Sleep Medicine; Department of Medicine; University of Pittsburgh School of Medicine; Pittsburgh, PA, USA; Department of Critical Care Medicine, Mercy St. Louis, St. Louis, MO, USA; Department of Emergency Medicine, Weill Cornell Medicine, New York, NY, USA; Department of Biobehavioral and Nursing Science, College of Nursing, University of South Carolina, Columbia, SC, USA; Department of Emergency Medicine, University of Iowa, Iowa City, IA, USA

**Author notes:** Corresponding author information: Chukwudi Onyemekwu, 3459 Fifth Avenue, Pittsburgh, PA 15213.

**Keywords:** Delirium, electroencephalography, machine learning algorithm, point-of-care EEG, intensive care unit

## Abstract

**Purpose:** Delirium affects around 40% of intensive care unit (ICU) patients and adversely affects survival, length of stay, and cognitive recovery. Despite this, implementation of systematic clinical delirium assessments is variable. We sought to validate a point-of-care electroencephalography (EEG)-based system (Ceribell, Inc.) for the detection of delirium in the ICU.

**Methods:** In this multicenter, prospective cohort study, we monitored adult ICU patients with the Ceribell EEG system for 6-8 hours for a minimum of three days. Up to three times per day, expert clinicians assessed participants for delirium and research coordinators completed Confusion Assessment Method for the ICU (CAM-ICU) assessments, which we used to calculate CAM-ICU-7 severity scores. We compared expert clinician diagnoses with Ceribell EEG algorithm output using data from 30-minute periods surrounding each clinician assessment.

**Results:** Among 225 participants analyzed (median age: 64 years, 60% male, 21% mechanically ventilated), expert clinicians performed 857 assessments and found 24% of participants were delirious during at least one assessment. The Ceribell EEG system detected delirium with a sensitivity of 81% (95% CI: 76% to 87%) and specificity of 81% (95% CI: 78% to 84%). The continuous Ceribell EEG system score correlated with CAM-ICU-7 delirium severity scores (bootstrapped median rho 0.58, IQR: 0.54-0.62, p <0.001).

**Conclusion:** In a mixed adult ICU population, the Ceribell EEG system identified delirium with good sensitivity and specificity. Its continuous output was associated with delirium severity, highlighting its potential ability to characterize underlying physiological patterns across the clinical delirium spectrum.

## Introduction

Delirium is common and harmful during critical illness, contributing to increased mortality, prolonged mechanical ventilation, delayed discharge from the intensive care unit (ICU) and hospital, and long-term cognitive impairment among survivors [1-5]. Though 20%-40% of critically ill patients and up to 80% of mechanically ventilated ICU patients develop delirium [6, 7], this common form of acute brain dysfunction goes unrecognized by many clinicians [8, 9].

The Confusion Assessment Method for the ICU (CAM-ICU) [10] and Intensive Care Delirium Screening Checklist (ICDSC) [11], which were developed to bridge this gap, are both sensitive and specific when used by trained ICU team members, but use of these tools in clinical settings is variable [12]. Barriers to delirium detection in the ICU include inadequate clinician education and confidence with screening tools, perceived complexity of the tools, competing clinical demands and nursing workload, and organizational culture [13, 14]. In addition, even when the CAM-ICU or ICDSC is reliably used, the waxing and waning of delirium features may mean that clinical assessments miss important changes.

Recent studies have shown that electroencephalography (EEG) can be used to detect delirium with good discrimination [15-17]. Limitations in conventional EEG, which usually requires a trained technologist for electrode placement and expert neurologist interpretation, make it unsuitable as a routine screening instrument, so EEG has not been widely used as a diagnostic tool for delirium in the ICU. Ceribell EEG (Ceribell, Inc., Sunnyvale, CA) is an EEG system that uses a portable 10-electrode headband and an artificial intelligence (AI)-assisted algorithm to record and analyze brain activity. The system, which accurately monitors seizure burden and is United States Food and Drug Administration (FDA)-cleared for seizure detection [18, 19], has the potential to address some limitations of clinical delirium monitoring.

We conducted a multicenter prospective cohort study to validate the performance of a novel EEG-based delirium detection algorithm in adult ICU patients. We hypothesized that the Ceribell EEG system has good discriminative ability for detecting delirium, using expert clinician assessments as the reference standard.

## Methods

### Study Design and Population

We completed a multicenter prospective cohort study recruiting adults (age ≥18 years) admitted to the ICU and who were fluent in English to complete the delirium assessments. We excluded patients who had a condition that prevented the use of the Ceribell EEG headband, including anatomical issues (e.g., hemicraniectomy with absent bone flap where EEG electrodes would be placed) or who were expected to need clinical continuous EEG monitoring during the proposed study period. We recruited in seven United States (US) hospitals, but only six completed expert clinician assessments: Mercy Hospital St. Louis, Naples Comprehensive Health Baker Hospital (NCH Baker Hospital), Stanford University Medical Center (SUMC), University of Iowa Health Care Medical Center, University of North Carolina (UNC) REX Hospital, and UPMC Presbyterian. The WCG Institutional Review Board (IRB) approved the study and served as the IRB in sites with a reliance agreement (UNC and University of Iowa). Local IRBs approved the study at all other sites. The study was exempt from FDA Investigational Device Exemptions (IDE) requirements and was registered at ClinicalTrials.gov (NCT04962815). This study was conducted in accordance with the ethical standards of the institutional committee and the 1964 Declaration of Helsinki and its later amendments.

### Study Procedures

After obtaining informed consent from the participant or an authorized representative, we monitored participants with the continuous Ceribell EEG system for up to 8 hours per day for a minimum of three days or until death or ICU discharge. During study days, trained research coordinators placed the Ceribell EEG headband on participants in the morning, connecting it to the recorder and ensuring proper electrode connection. They visited participants several hours later to check electrode connection and, if needed, troubleshoot electrode connection and assess for skin irritation. They removed the Ceribell EEG headband in the evening, performing a final skin assessment. Research coordinators assessed participants three times each study day with the CAM-ICU [10] if the participant’s Richmond Agitation Sedation Scale (RASS) [20] was ≥-3. For each CAM-ICU assessment, the number of errors was recorded for each feature to calculate the CAM-ICU 7. At least once daily and sometimes more often, depending on availability, a qualified clinician completed a delirium assessment within 20 minutes of the research coordinator’s CAM-ICU assessment. We instructed qualified clinicians to use all information available to identify delirium according to established clinical criteria [21]. These included bedside assessments, physical exams, medical record review, and information about the clinical context, e.g., obtained from the bedside nurse or other members of the ICU team.

### EEG Algorithm

Ceribell developed the EEG-based delirium monitoring algorithm using a dataset of 267 adult ICU patients, including a cohort of 47 participants from the current study set aside for development, all of whom were monitored with the Ceribell EEG system. In this development dataset, delirium was identified in 25% of the patients using either CAM-ICU assessment or clinician’s diagnosis. The development dataset was split into a training (80%) and a testing cohort (20%). To evaluate the algorithm, 30-minute windows of recording time around the assessment or diagnosis were used to generate a mean of the algorithm output. This mean was then binarized based on a prespecified threshold to produce a binary algorithm output indicating presence or absence of delirium. In the testing cohort, the algorithm had a sensitivity of 89% and a specificity of 90% for delirium when using clinical assessment as reference standard, with an area under the curve of 0.96.

### Algorithm Validation

To ensure unbiased evaluation, we evaluated the performance of the algorithm in a validation dataset that did not contain any participants included in the development dataset. In this validation dataset, we validated the algorithm’s continuous output against expert clinicians’ assessments. To evaluate algorithmic performance against this reference standard, we extracted 30-minute window of Ceribell EEG data, centered around the timestamps of each clinical assessment to generate the corresponding model output. When the reference standard clinical delirium assessment was performed at the edges of the EEG recording (i.e., less than 15 minutes from the beginning or end of the EEG recording), the monitoring window was shifted such that 30 minutes of EEG data were always included.

### statistical Analysis

Using expert clinician assessments as the reference standard, we derived true-positive, false-positive, false-negative, and true-negative data for the Ceribell EEG Delirium algorithm output. We then calculated the sensitivity and specificity, together with the corresponding 95% confidence intervals, at the assessment level. Since participants underwent multiple assessments, we accounted for within-patient correlation using the bias-correcting accelerated bootstrap method when calculating the 95% confidence interval.

In addition to comparing the Ceribell EEG Delirium algorithm’s binary diagnosis with that of expert clinicians, we explored the relationship between the mean algorithm’s output and delirium severity as assessed by the CAM-ICU-7 [22] in a subset of 174 participants with assessments during which the individual features of the CAM-ICU were recorded. First, we categorized participant-assessments as no delirium (CAM-ICU-7: 0), subsyndromal delirium (CAM-ICU-7: 1-2), mild-to-moderate delirium (CAM-ICU-7: 3-5), or severe delirium (CAM-ICU-7: 6-7). Then, we used a linear mixed-effects model to estimate the change in Ceribell EEG Delirium algorithm score associated with each delirium severity category, using no delirium as the reference category. We adjusted for unbalanced repeated measures per participant and time difference between assessments. We also included the time from recording start to start of the assessment as a fixed effect and participant as a random effect.

Lastly, we used Spearman’s rank correlation to assess the correlation between the Ceribell EEG Delirium mean algorithm’s output and the raw CAM-ICU-7 score. To account for within-patient correlation, we used bootstrapping, randomly selecting a single assessment per participant in each of 5,000 iterations, reporting the median and 25th and 75th quartiles of the Spearman rho and using a permutation test to estimate the exact p-value.

## Results

From July 2022 to September 2025, we recruited 495 patients. As shown in **Figure 1**, 207 eligible patients were set aside for current and future development datasets. Of the 288 remaining patients, 5 (1.7%) met at least one exclusion criterion, 27 (9.3%) were enrolled but discharged from the ICU before any EEG recording or assessments were done, 16 (5%) were from a single site that ran an initial version of the protocol that did not collect any clinician’s diagnosis and the site was closed after protocol changes, and 15 (5%) had poor EEG signal quality or data-entry issues. A total of 225 participants who underwent 857 delirium assessments were included for the algorithm validation analysis.

**Figure 1.**
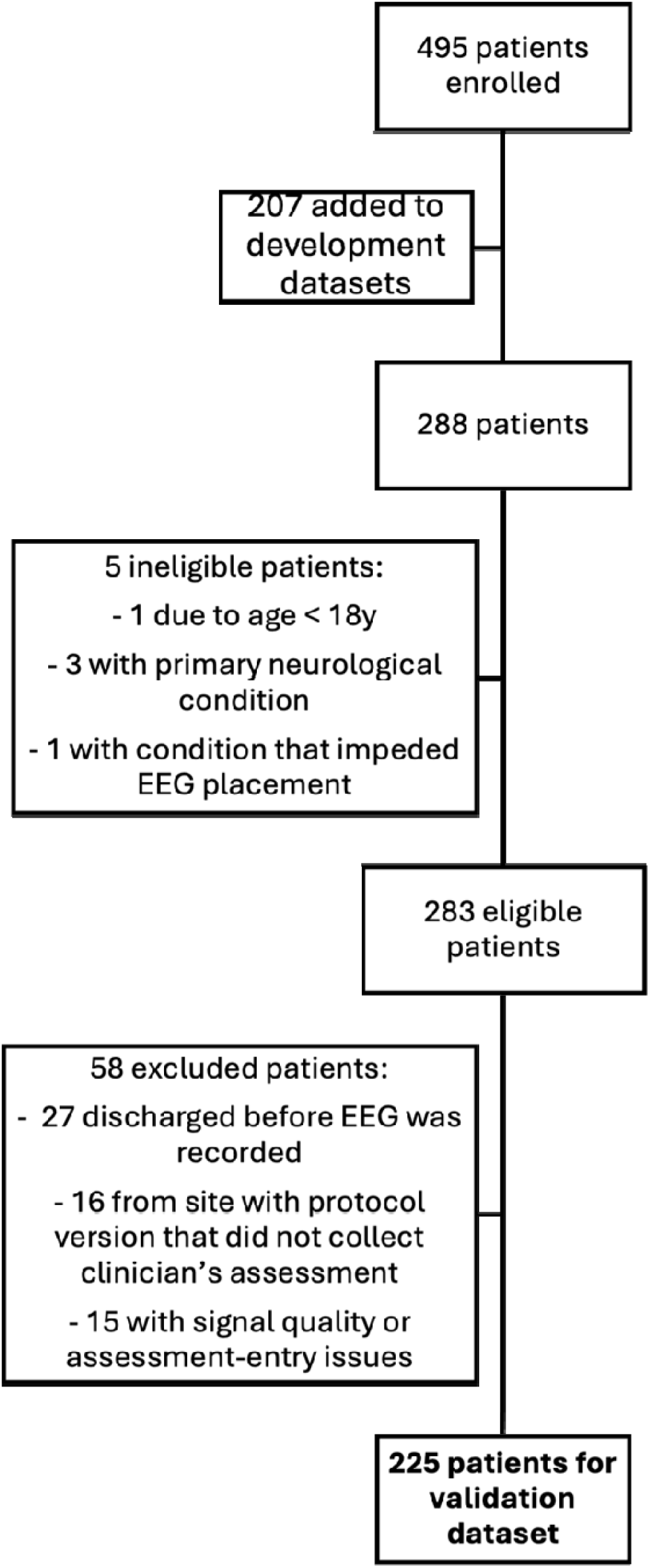
Validation dataset patient enrollment, exclusion, and inclusion for analysis.

The study population was similar to general medical ICU populations: median age was 64 years, 60% of patients were male, and 21% were receiving invasive mechanical ventilation (**Table 1**). The median recording duration was 4.8 h (IQR: 2.8-5.8), the median number of clinician assessments per patient was 3 (IQR: 2-5), and median number of unique monitoring days was 1 (IQR: 1-2). The expert clinician assessments determined that 54 (24%) participants were delirious at some point during the study. These participants were delirious during 205 expert clinician assessments; RASS was <0 (i.e., delirium was hypoactive) during 172 (84%) of these assessments.

**Table 1.** Validation dataset demographics.

| Characteristic | Overall<sup>1</sup><br>(N = 225) | Positive<br>Clinical<br>Diagnosis<sup>1</sup><br>(N = 54) | Negative<br>Clinical<br>Diagnosis<sup>1</sup><br>(N = 171) |
| --- | --- | --- | --- |
| Age (years) | 64 (56, 73) | 66 (56, 70) | 64 (55, 73) |
| Sex (Female) | 90 (40%) | 19 (35%) | 71 (42%) |
| Mechanically Ventilated | 47 (21%) | 25 (46%) | 22 (13%) |
| Number of clinician's assessments<br>per patient | 3 (2, 5) | 5 (3, 8) | 3 (2, 5) |
| EEG Recording duration (hours) | 4.8 (2.8, 5.8)□ | 5.4 (3.8, 6.0) | 4.2 (2.4, 5.6) |
| **Admission Diagnosis Groups<sup>2</sup>** |  |  |  |
| Acute Respiratory Failure | 21 (9.3%) | 6 (11.1%) | 14 (8.2%) |
| Sepsis | 19 (8.4%) | 4 (7.4%) | 15 (8.8%) |
| Stroke | 18 (8.0%) | 7 (13%) | 11 (6.4%) |
| Septic Shock | 16 (7.1%) | 2 (3.7%) | 14 (8.2%) |
| Shock | 13 (5.8%) | 3 (5.6%) | 10 (5.8%) |
| Cardiovascular | 11 (4.9%) | 2 (3.7%) | 9 (4.7%) |
| GI Bleed/Ischemia | 7 (3.2%) | 1 (1.8%) | 8 (4.7%) |
| Decompensation of Hematologic<br>Disease | 7 (3.1%) | 0 | 7 (4.1%) |
| Infection | 7 (3.1%) | 1 (1.8%) | 6 (3.5%) |
| Pneumonia | 7 (3.1%) | 2 (3.7%) | 5 (2.9%) |
| Toxic Encephalopathy | 7 (3.1%) | 2 (3.7%) | 5 (2.9%) |
| Others | 76 (33.8%) | 16 (29.6%) | 60 (35%) |
| Unknown | 14 | 5 | 9 |

<sup>1</sup>Median (Q1, Q3); N (%)
<sup>2</sup>Wilcoxon rank-sum test; Fisher's exact test; Chi-squared test
<sup>3</sup>Listing top eleven grouped diagnoses, which were present at ICU admission but not necessarily still present during the study.
CAM-ICU: Confusion Assessment Method for the Intensive Care Unit; ICU: intensive care unit; TIA: transient ischemic attack; SAH: subarachnoid hemorrhage; COPD: chronic-obstructive pulmonary disease

The Ceribell EEG Delirium system demonstrated good sensitivity and specificity, capturing 167 of 205 positive clinical delirium assessments, with a sensitivity of 81% (95% CI: 76%-87%) and specificity of 81% (95% CI: 78%-84%). The full 2×2 cross-tabulation of the Ceribell performance versus expert clinician diagnosis is provided in **Online Resource 1**. Illustrative examples of multiday Ceribell EEG Delirium system output and expert clinician assessments are shown in **Figure 2**, to exemplify the system’s ability to capture day-to-day fluctuations in participant’s mental states.

**Figure 2.**
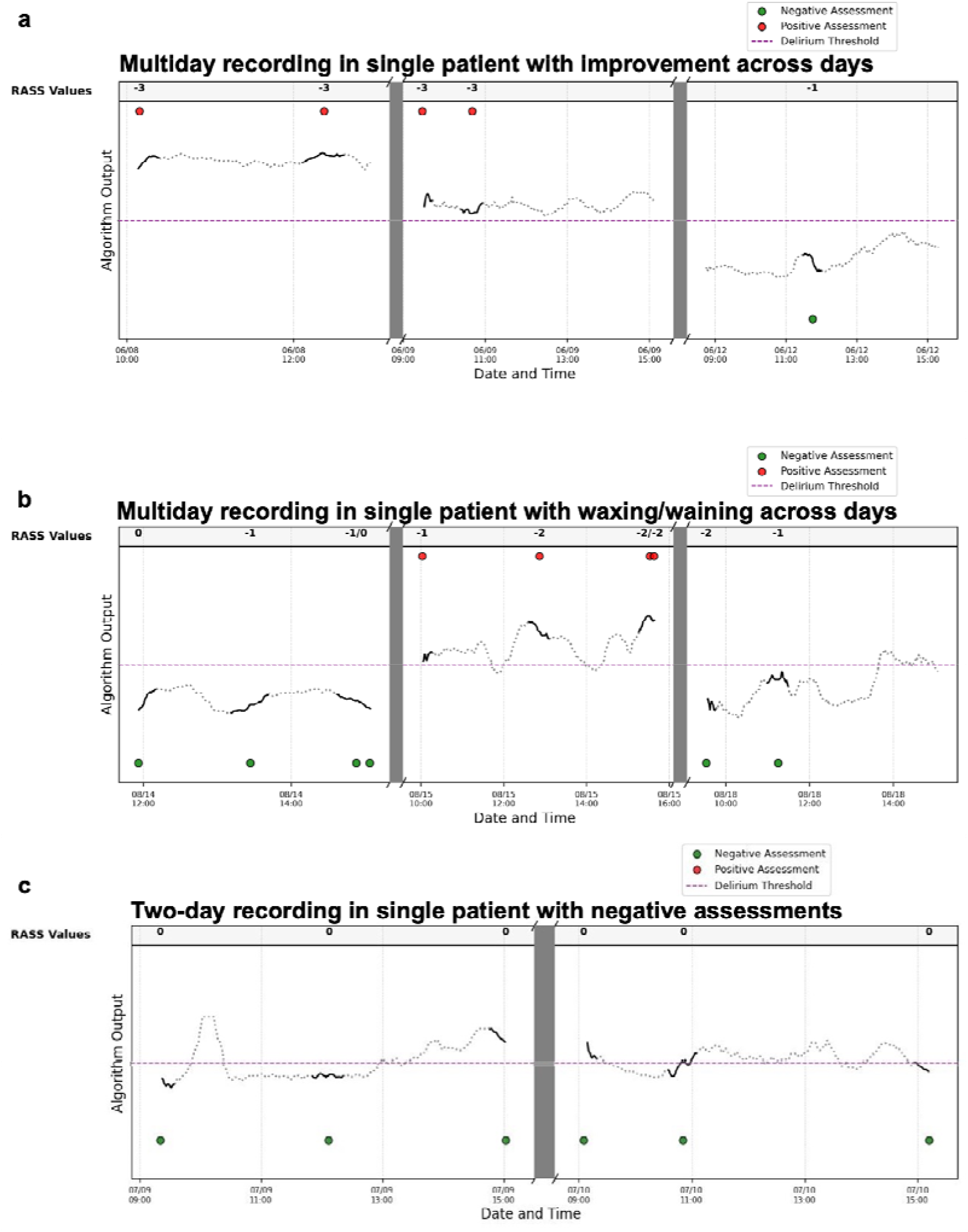
Exemplar cases of multiday algorithm output with diagnoses from clinical assessments. The time series shows the continuous algorithm output (black solid and dotted lines) over multiday ICU stays for individual participants. Each solid line represents data within 30 minutes of the clinical assessment, and the dotted line shows the output at every other point of the recording. The purple dashed line shows the threshold for the algorithm to detect delirium. Circular markers show the binary clinical assessments (red = positive delirium, green = negative delirium). Grayed areas illustrate breaks in the recording. RASS sedation scores at the time of the clinical assessments are shown at the top. **a)** Multiday recording with true-positive across three sequential days of recording for one participant. **b)** In a different participant, three-day recording with fluctuations in delirium clinical assessment. **c)** A third participant with a two-day recording with false-positive per the clinical assessment, but w observe the scores were close to the threshold, with fluctuations throughout the day.

In an exploratory analysis, we found that the mean continuous Ceribell EEG Delirium score differed significantly by CAM-ICU-7 delirium severity category (**Figure 3**). Using no delirium as the reference category, the mean Ceribell EEG Delirium score was 5.5 (95% C.I.: 1.9-8.9) points higher during subsyndromal delirium, 18.3 (95% C.I.: 14.2-22.3) points higher during mild-to-moderate delirium, and 25.4 (95% C.I.: 20.8-30.0) points higher during severe delirium. As shown in **Figure 4**, the continuous Ceribell EEG Delirium score was moderately correlated with the CAM-ICU-7 (median rho 0.58, interquartile range 0.54-0.62; p <0.001).

**Figure 3.**
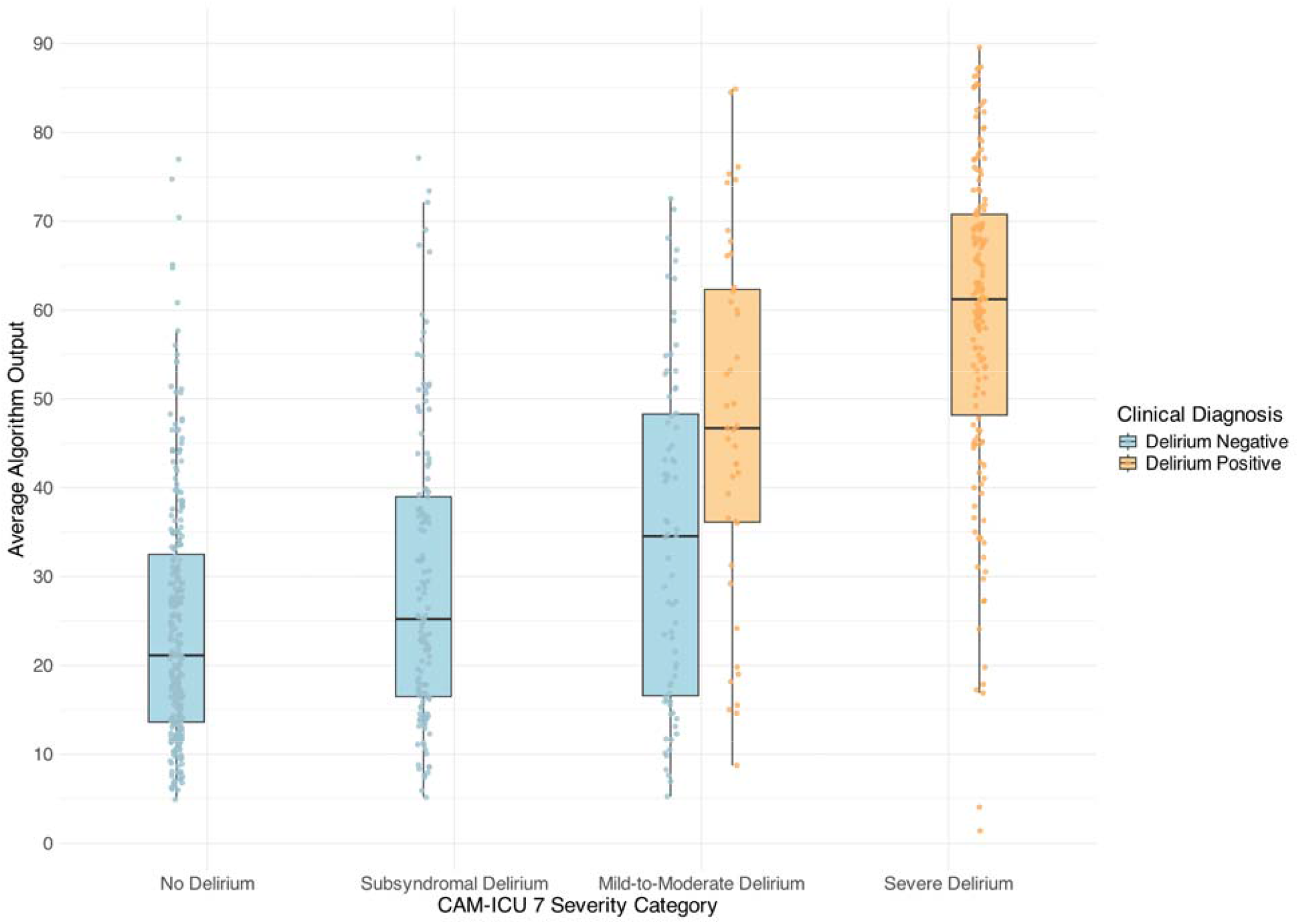
Association between delirium severity categories, per CAM-ICU 7 score, and the Ceribell delirium algorithm output. Each category in the x-axis shows boxplot interquartile range of the algorithm output (y-axis); the dots represent individual assessments within each group. Blue and yellow colors further categorize the assessments that were negative and positive per the clinical diagnosis, respectively.

**Figure 4.**
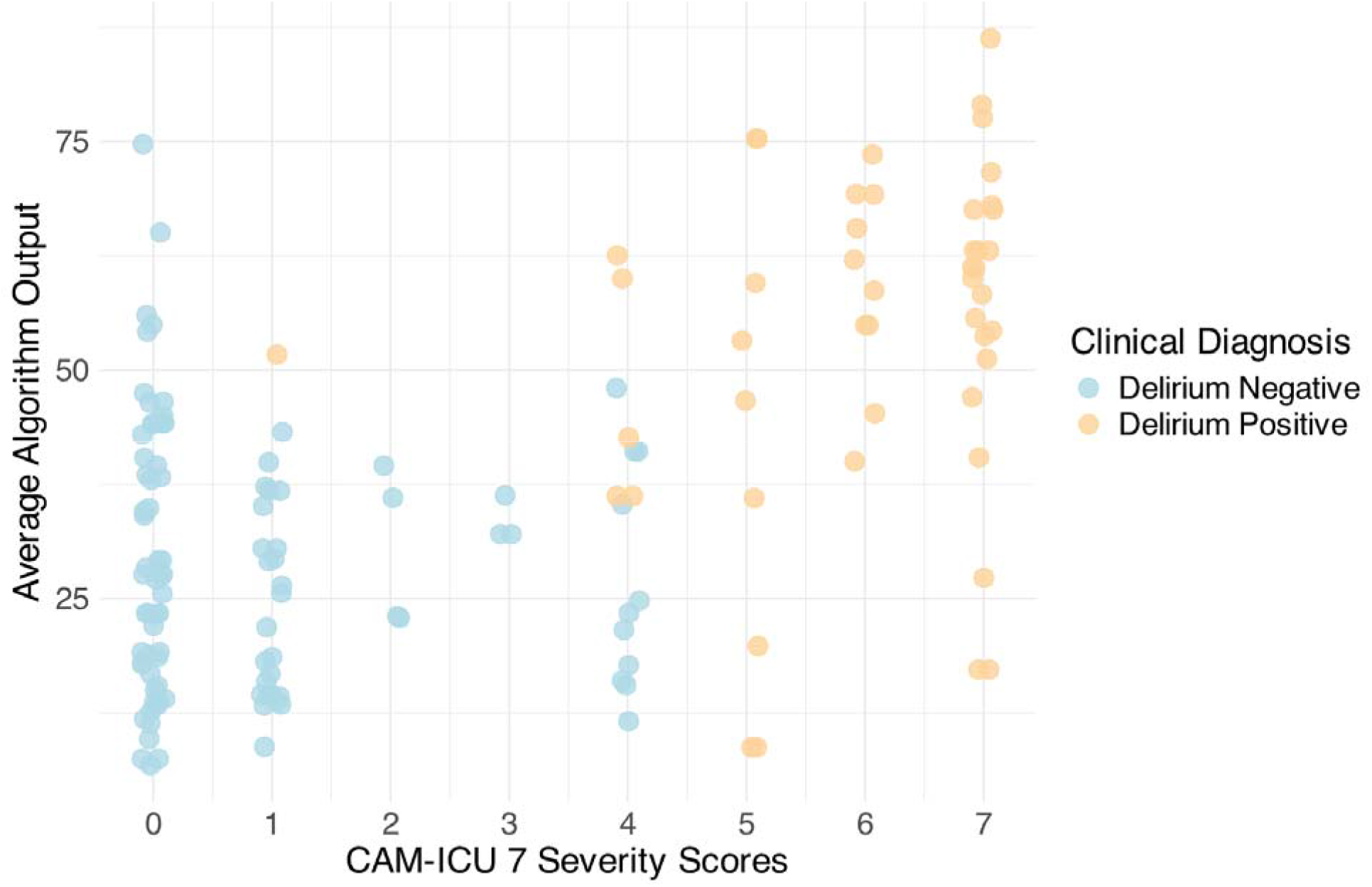
CAM-ICU 7 severity scores across 172 participants with errors and individual features data. If the participant had multiple assessments throughout the study, a single assessment was chosen at random with the corresponding algorithm output. Jitter was added in the x-axis to each datapoint (width = 0.1). The markers color represents whether the participant had a positive (light orange) or negative (light blue) clinical diagnosis corresponding to the plotted assessment.

## Discussion

In this large, multicenter, prospective study, we found that the portable Ceribell EEG system was both sensitive and specific for delirium in the ICU when using delirium diagnoses made by expert clinicians as the reference standard. The study cohort was heterogeneous and included mechanically ventilated patients, many of whom were sedated. These results suggest that the Ceribell EEG system, which recently received US FDA 510(k) clearance for delirium monitoring, can be used to support reliable and continuous detection of delirium in ICUs where clinical assessments are infrequent and sometimes not reliably completed.

EEG provides an objective measure of brain activity that can be recorded continuously, a feature that may be advantageous when detecting delirium given that fluctuation is so common during critical illness. EEG-based monitors have the potential to continuously detect changes in brain activity and may thereby alert the ICU team about deteriorating brain function earlier than conventional clinical assessments. Indeed, in an exploratory analysis, we found the Ceribell EEG Delirium algorithm score correlated with the CAM-ICU-7. This continuous output of the algorithm could facilitate delirium detection earlier than binary point-in-time assessments. In turn, earlier, reliable detection of worsening delirium could alter management and support implementation of delirium prevention measures, a hypothesis that should be tested in future clinical trials.

As shown in examples from our study, the Ceribell EEG system can assess EEG data over longer periods of time than are assessed during one, or even multiple, clinical assessments. Figure 2 shows that some participants’ Ceribell EEG scores crossed the threshold for delirium multiple times in a day. Additionally, though this hypothesis could not be tested in the current study, it is possible that the delirium assessments currently classified as false positives (i.e., the Ceribell EEG Delirium algorithm score was above the classification threshold but the expert clinician did not diagnose delirium) represent meaningful instances of acute brain dysfunction detected by the EEG despite the inability of an expert clinician to detect delirium.

Future research should evaluate whether EEG-detected changes in brain function during critical illness are clinically relevant even if they are not detected by clinical delirium assessments.

Several previous studies of point-of-care EEG-based delirium monitors were single-center proof-of-concept studies limited by small sample sizes [23-27]. Other studies included only specific populations, such as older adults [27, 28], psychiatry consultation referrals [23], or post-operative patients [26, 29], Finally, many prior studies relied on a single EEG snapshot rather than continuous, repeated recordings [23, 28, 30]. In contrast, our current study included a large, mixed ICU patient population recruited in multiple hospitals, multiple reference standard delirium assessments per participant, and use of a portable device that continuously records EEG activity, is commercially available, and is now FDA-cleared for detection of delirium with good sensitivity and specificity in the ICU.

Our study has several strengths. First, reference-rater clinicians completed up to three assessments per day to best capture the presence of delirium, and found that the prevalence of delirium in our cohort was similar to that reported in the literature [6, 7]. Second, we captured over 800 assessments across 225 unique participants in a mixed ICU cohort, making it one of the largest studies of an EEG-based delirium monitor. Third, we used as our reference standard the diagnoses of expert clinicians who were trained in delirium diagnosis and who relied on all available information about the participant’s condition. Finally, we also calculated the CAM-ICU-7, which enabled a more granular assessment of delirium severity, and in an exploratory analysis found that the Ceribell EEG Delirium algorithm score correlates with this measure of delirium severity.

Our study also had several limitations. First, despite being larger than previous similar studies, our study was not large enough to evaluate how the Ceribell EEG system performs in important subgroups (e.g., hypoactive delirium, sedative-associated delirium). Sedation, in particular, can induce EEG changes that may be separate from those reflecting delirium. Future research is needed to assess discrimination of delirium in lightly or moderately sedated patients. Additionally, though we sought to complete assessments in participants with both hypoactive and hyperactive delirium, it is possible that hyperactive delirium, which was uncommon during our study assessments, causes data quality issues, making EEG data less reliable during agitated delirium. Third, the recordings were around 5 hours in duration during the day up to the early evening shifts, the system may need further evaluation during long-term monitoring.

Finally, we did not evaluate short- or long-term outcomes to determine whether Ceribell EEG-detected delirium predicts adverse outcomes as does clinically detected delirium. Finally, this study examined only detection of delirium and did not evaluate the effects of a patient management strategy involving the Ceribell EEG system.

## Conclusion

In this large, multicenter study of a mixed adult ICU population, we found the Ceribell EEG system detected delirium with good sensitivity and specificity, and the continuous algorithm output correlated with CAM-ICU-7 severity score. This system has the potential to address multiple barriers to reliable, accurate delirium detection in the ICU, thereby improving delirium diagnosis, documentation, and management in the ICU. Future studies are needed to examine performance in specific clinical subgroups and to examine the effects of incorporating the Ceribell EEG system into clinical workflows on clinical outcomes.

### Take-Home Message

Delirium detection in the ICU relies on intermittent clinical assessments, leaving gaps in monitoring. This multicenter prospective study demonstrates that a point-of-care EEG system (Ceribell) detects delirium with good sensitivity (81%) and specificity (81%) against expert clinical diagnosis. These findings support point-of-care EEG as a feasible, objective, continuous adjunct for delirium surveillance in critically ill adults, complementing standard intermittent behavioral assessments like CAM-ICU.

## Data Availability

The data produced in this study are not publicly available, and we do not have permission to share the raw data under the terms of the IRB determination. The reviews generated for this study are the property of Ceribell, Inc., but reasonable requests may be considered by the corresponding author.

## Statements and Declarations

## Acknowledgements

We thank and acknowledge Dr. Viday Patel, who served as site investigator at Naples Hospital, and Dr. Jose Maldonado, who served as site investigator at Stanford University Medical Center. Guarantor: Chukwudi A. Onyemekwu (C.A.O.)

## Author Contributions

C.A.O., M.D.M., T.S., N.T.P., K.M.T., F.S., and T.D.G. designed, collected, analyzed, and interpreted the data. S.K., M.A.S, X.G., G.T., and T.D.G. performed critical data quality assurance and data analysis. C.A.O., S.K., M.A.S., and T.D.G. drafted and revised the manuscript. N.T.P., K.M.T., M.D.M., S.L., B.K., M.A.M., F.S., T.S., and T.D.G. performed critical reviews and edits of the manuscript in preparation for publication.

## Consent to participate

Informed consent was obtained from all individual participants or authorized legal representatives prior to participation in the study.

## References

1. Devlin JW, Skrobik Y, Gélinas C, Needham DM, Slooter AJC, Pandharipande PP, Watson PL, Weinhouse GL, Nunnally ME, Rochwerg B et al: Clinical Practice Guidelines for the Prevention and Management of Pain, Agitation/Sedation, Delirium, Immobility, and Sleep Disruption in Adult Patients in the ICU. Critical Care Medicine 2018, 46(9):e825–e873.

2. Shehabi Y, Riker RR, Bokesch PM, Wisemandle W, Shintani A, Ely EW: Delirium duration and mortality in lightly sedated, mechanically ventilated intensive care patients. Crit Care Med 2010, 38(12):2311–2318.

3. Hughes CG, Hayhurst CJ, Pandharipande PP, Shotwell MS, Feng X, Wilson JE, Brummel NE, Girard TD, Jackson JC, Ely EW et al: Association of delirium during critical illness with mortality: Multicenter prospective cohort study. Anesth Analg 2021, 133(5):1152–1161.

4. Girard TD, Jackson JC, Pandharipande PP, Pun BT, Thompson JL, Shintani AK, Gordon SM, Canonico AE, Dittus RS, Bernard GR et al: Delirium as a predictor of long-term cognitive impairment in survivors of critical illness. Crit Care Med 2010, 38(7):1513–1520.

5. Pandharipande PP, Girard TD, Jackson JC, Morandi A, Thompson JL, Pun BT, Brummel NE, Hughes CG, Vasilevskis EE, Shintani AK et al: Long-term cognitive impairment after critical illness. N Engl J Med 2013, 369(14):1306–1316.

6. Krewulak KD, Stelfox HT, Leigh JP, Ely EW, Fiest KM: Incidence and Prevalence of Delirium Subtypes in an Adult ICU: A Systematic Review and Meta-Analysis*. Critical Care Medicine 2018, 46(12):2029–2035.

7. Gibb K, Seeley A, Quinn T, Siddiqi N, Shenkin S, Rockwood K, Davis D: The consistent burden in published estimates of delirium occurrence in medical inpatients over four decades: a systematic review and meta-analysis study. Age and Ageing 2020, 49(3):352–360.

8. van Eijk MM, van Marum RJ, Klijn IA, de Wit N, Kesecioglu J, Slooter AJ: Comparison of delirium assessment tools in a mixed intensive care unit. Crit Care Med 2009, 37(6):1881–1885.

9. Chanques G, Ely EW, Garnier O, Perrigault F, Eloi A, Carr J, Rowan CM, Prades A, de Jong A, Moritz-Gasser S et al: The 2014 updated version of the Confusion Assessment Method for the Intensive Care Unit compared to the 5th version of the Diagnostic and Statistical Manual of Mental Disorders and other current methods used by intensivists. Ann Intensive Care 2018, 8(1):33.

10. Ely EW, Inouye SK, Bernard GR, Gordon S, Francis J, May L, Truman B, Speroff T, Gautam S, Margolin R et al: Delirium in mechanically ventilated patients: validity and reliability of the confusion assessment method for the intensive care unit (CAM-ICU). JAMA 2001, 286(21):2703–2710.

11. Bergeron N, Dubois MJ, Dumont M, Dial S, Skrobik Y: Intensive Care Delirium Screening Checklist: evaluation of a new screening tool. Intensive Care Med 2001, 27(5):859–864.

12. Titlestad I, Haugarvoll K, Solvang S-EH, Norekvål TM, Skogseth RE, Andreassen OA, Årsland D, Neerland BE, Nordrehaug JE, Tell GS et al: Delirium is frequently underdiagnosed among older hospitalised patients despite available information in hospital medical records. Age and Ageing 2024, 53(2).

13. Ragheb J, Norcott A, Benn L, Shah N, McKinney A, Min L, Vlisides PE: Barriers to delirium screening and management during hospital admission: a qualitative analysis of inpatient nursing perspectives. BMC Health Services Research 2023, 23(1):712.

14. Hook ML, Lindroth HL, McGuire D, Reopelle SL, Strand SA, Jacobson N: Factors Affecting Nurses’ Ability to Identify Delirium in Acute Care Settings. Res Gerontol Nurs 2026:1–13.

15. Kimchi EY, Neelagiri A, Whitt W, Sagi AR, Ryan SL, Gadbois G, Groothuysen D, Westover MB: Clinical EEG slowing correlates with delirium severity and predicts poor clinical outcomes. Neurology 2019, 93(13):e1260–e1271.

16. Boord MS, Moezzi B, Davis D, Ross TJ, Coussens S, Psaltis PJ, Bourke A, Keage HAD: Investigating how electroencephalogram measures associate with delirium: A systematic review. Clin Neurophysiol 2021, 132(1):246–257.

17. Williams Roberson S, Azeez NA, Fulton JN, Zhang KC, Lee AXT, Ye F, Pandharipande P, Brummel NE, Patel MB, Ely EW: Quantitative EEG signatures of delirium and coma in mechanically ventilated ICU patients. Clin Neurophysiol 2023, 146:40–48.

18. Vespa PM, Olson DM, John S, Hobbs KS, Gururangan K, Nie K, Desai MJ, Markert M, Parvizi J, Bleck TP et al: Evaluating the Clinical Impact of Rapid Response Electroencephalography: The DECIDE Multicenter Prospective Observational Clinical Study*. Critical Care Medicine 2020, 48(9).

19. Kamousi B, Karunakaran S, Gururangan K, Markert M, Decker B, Khankhanian P, Mainardi L, Quinn J, Woo R, Parvizi J: Monitoring the Burden of Seizures and Highly Epileptiform Patterns in Critical Care with a Novel Machine Learning Method. Neurocritical Care 2021, 34(3):908–917.

20. Sessler CN, Gosnell MS, Grap MJ, Brophy GM, O’Neal PV, Keane KA, Tesoro EP, Elswick RK: The Richmond Agitation-Sedation Scale: validity and reliability in adult intensive care unit patients. Am J Respir Crit Care Med 2002, 166(10):1338–1344.

21. American Psychiatric Association., American Psychiatric Association. DSM-5 Task Force.: Diagnostic and statistical manual of mental disorders : DSM-5, 5th edn. Washington, D.C.: American Psychiatric Association; 2013.

22. Khan BA, Perkins AJ, Gao S, Hui SL, Campbell NL, Farber MO, Chlan LL, Boustani MA: The Confusion Assessment Method for the ICU-7 Delirium Severity Scale: A Novel Delirium Severity Instrument for Use in the ICU. Critical Care Medicine 2017, 45(5).

23. Luo A, Muraida S, Pinchotti D, Richardson E, Ye E, Hollingsworth B, Win A, Myers O, Langsjoen J, Valles E et al: Bispectral Index Monitoring With Density Spectral Array for Delirium Detection. J Acad Consult Liaison Psychiatry 2021, 62(3):318–329.

24. Mulkey MA, Gantt LT, Hardin SR, Munro CL, Everhart DE, Kim S, Schoeman AM, Roberson DW, McAuliffe M, Olson DM: Rapid Handheld Continuous Electroencephalogram (EEG) Has the Potential to Detect Delirium in Older Adults. Dimens Crit Care Nurs 2022, 41(1):29–35.

25. Mulkey MA, Huang H, Albanese T, Kim S, Yang B: Supervised deep learning with vision transformer predicts delirium using limited lead EEG. Sci Rep 2023, 13(1):7890.

26. Guo Z, Wan W, Liu W, Liu L, Yang Y, Yang C, Cui X: Quantitative electroencephalography predicts postoperative delirium in adult cardiac surgical patients from a prospective observational study. Sci Rep 2024, 14(1):31101.

27. Shinozaki G, Chan AC, Sparr NA, Zarei K, Gaul LN, Heinzman JT, Robles J, Yuki K, Chronis TJ, Ando T et al: Delirium detection by a novel bispectral electroencephalography device in general hospital. Psychiatry Clin Neurosci 2018, 72(12):856–863.

28. Yamanashi T, Kajitani M, Iwata M, Crutchley KJ, Marra P, Malicoat JR, Williams JC, Leyden LR, Long H, Lo D et al: Topological data analysis (TDA) enhances bispectral EEG (BSEEG) algorithm for detection of delirium. Sci Rep 2021, 11(1):304.

29. Numan T, van den Boogaard M, Kamper AM, Rood PJT, Peelen LM, Slooter AJC, Abawi M, van den Boogaard M, Claassen JAHR, Coesmans M et al: Delirium detection using relative delta power based on 1-minute single-channel EEG: a multicentre study. British Journal of Anaesthesia 2019, 122(1):60–68.

30. Ditzel FL, Hut SCA, van den Boogaard M, Boonstra M, Leijten FSS, Wils E-J, van Nesselrooij T, Kromkamp M, Rood PJT, Röder C et al: DeltaScan for the Assessment of Acute Encephalopathy and Delirium in ICU and non-ICU Patients, a Prospective Cross-Sectional Multicenter Validation Study. The American Journal of Geriatric Psychiatry 2024, 32(9):1093–1104.

