## Supplemental Table 1. for "Validation of an EEG-based Delirium Monitor in the ICU"

**Online Resource 1.**

2X2 Cross-tabulation of expert clinician assessment versus Ceribell algorithm performance of 225 patients at the assessment level

| **N = 857 assessments** | **Expert Clinician Assessment Delirium (+)** | **Expert Clinician Assessment Delirium (-)** |
|---|---|---|
| **Ceribell (+)** | 167 | 126 |
| **Ceribell (-)** | 38 | 526 |
